# Statistical Analysis Plan for the Helmet Non-Invasive Ventilation for COVID-19 Patients (Helmet-COVID) Randomized Controlled Trial

**DOI:** 10.1101/2021.07.26.21260421

**Authors:** Yaseen Arabi, Haytham Tlayjeh, Sara Aldekhyl, Hasan Al-Dorzi, Sheryl Ann Abdukahil, Jesna Jose, Mohammad Khulaif Al Harbi, Husain Al Haji, Mohammed Al Mutairi, Omar Al Zumai, Eman Al Qasim, Wedyan Al Wehaibi, Saad Al Qahtani, Mohammed Alshahrani, Talal Albrahim, Ahmed Mady, Ali Al Bshabshe, Zohair Al Aseri, Zainab Al Duhailib, Ayman Kharaba, Rakan Alqahtani, Haifa Algethamy, Omar Alfaris, Omar Alnafel, Abdulrahman A Al-Fares

**Affiliations:** Intensive Care Department, Ministry of National Guard Health Affairs, King Abdullah International Medical Research Center, King Saud Bin Abdulaziz University for Health Sciences, Riyadh, Saudi Arabia; College of Medicine, King Saud Bin Abdulaziz University for Health Sciences, King Abdullah International Medical Research Center, Ministry of National Guard Health Affairs, Riyadh, Saudi Arabia; Bioinformatics and Biostatistics Department, King Abdullah International Medical Research Center, King Saud Bin Abdulaziz University for Health Sciences, Ministry of National Guard Health Affairs, Riyadh, Saudi Arabia; Department of Anesthesia, Ministry of National Guard Health Affairs, King Abdullah International Medical Research Center, King Saud Bin Abdulaziz University for Health Sciences, Riyadh, Saudi Arabia; Respiratory Services Department, Ministry of National Guard Health Affairs, King Abdullah International Medical Research Center, King Saud Bin Abdulaziz University for Health Sciences, Riyadh, Saudi Arabia; Research Office, King Abdullah International Medical Research Center, King Saud Bin Abdulaziz University for Health Sciences, Riyadh, Saudi Arabia, Ministry of National Guard Health Affairs, Riyadh, Saudi Arabia; Department of Emergency and Critical Care, King Fahad Hospital of the University, Imam Abdulrahman Bin Faisal University, Al Khobar, Kingdom of Saudi Arabia; Department of Critical Care, King Fahad Hospital of the University, Imam Abdulrahman Bin Faisal University, Al Khobar, Kingdom of Saudi Arabia; Intensive Care Department, King Saud Medical City, Riyadh, Saudi Arabia, College of Medicine, Tanta University, Egypt; Department of Critical Care Medicine, King Khalid University, Aseer Central Hospital, Abha, Kingdom of Saudi Arabia; Emergency and Intensive Care Departments, College of Medicine, King Saud University, Riyadh, Saudi Arabia; Adult Critical Care Department, King Faisal Specialist Hospital & Research Center, Riyadh, Saudi Arabia; Pulmonary & Critical Care Departments, King Fahad Hospital Madinah, Critical Care Units- Madinah Region, Ministry of Health; Department of Critical Care, King Khalid University Hospital, King Saud University Medical City, Riyadh, Saudi Arabia; Department of Anesthesia, King Abdulaziz University Hospital, Jeddah, Saudi Arabia; Internal Medicine and Intensive Care Department, King Salman Specialist Hospital, Hail; Department of Anesthesia, Critical Care Medicine and Pain Medicine, Al-Amiri Hospital, Ministry of Health, Kuwait

**Author notes:** **Corresponding author Yaseen Arabi, MD FCCP, FCCM, ATSF [YA]** Intensive Care Department, Ministry of National Guard Health Affairs, Riyadh, Saudi Arabia, King Abdullah International Medical Research Center, King Saud Bin Abdulaziz University for Health Sciences, Riyadh, Saudi Arabia.

## Abstract

**Background and objective:** Noninvasive respiratory support is frequently needed for patients with acute hypoxemic respiratory failure due to coronavirus disease 19 (COVID-19). Helmet noninvasive ventilation having multiple advantages over other support modalities but data about effectiveness are limited.

**Methods:** In this multicenter randomized trial of helmet non-invasive ventilation for COVID-19 patients (Helmet-COVID), 320 adult ICU patients with suspected or confirmed COVID-19 and acute hypoxemic respiratory failure (with a ratio of arterial oxygen partial pressure to fraction (percent) of inspired oxygen (PaO_2_/FiO_2_) <200 despite supplemental oxygen with a partial/non-rebreathing mask at a flow rate >10 L/min or above) will be randomized to helmet-noninvasive ventilation with usual care or usual care alone. The primary outcome is death from any cause within 28 days after randomization. The trial has 80% power to detect a 15% absolute risk reduction from 40% to 25%.

**Conclusion:** Consistent with international guidelines, we developed a detailed plan to guide the analysis of the Helmet-COVID trial. This plan specifies the statistical methods for the evaluation of primary and secondary outcomes to facilitate unbiased analyses of clinical data.

**Trial registration:** Clinicaltrials.gov: NCT04477668 (registered on July 20, 2020)

## Introduction

Acute hypoxemic respiratory failure is a common feature of severe coronavirus disease 19 (COVID-19) and frequently requires respiratory support. As invasive mechanical ventilation carries high morbidity and mortality, other respiratory modalities, such as high-flow nasal oxygen and noninvasive ventilation (NIV) by face mask and helmet, have been suggested and increasingly practiced. Helmet NIV has multiple advantages over other modalities that may include more effective seal, less transmission of the virus, more effective delivery of positive end-expiratory pressure, and greater tolerance.(1) Helmet NIV has been investigated as a treatment in adult patients with acute hypoxic respiratory failure.(2-6) A network meta-analysis of 25 studies that included 3804 patients with acute hypoxemic respiratory failure for reasons other than COVID-19 found significant lower intubation (risk ratio, 0.26; 95% credible interval, 0.14-0.46) and mortality (risk ratio, 0.40; 95% credible interval, 0.24-0.63) risks with helmet NIV compared with standard oxygen therapy.(7) The absence of high-quality evidence on helmet NIV in COVID-19 led to the design and conduct of randomized controlled trials (RCTs). Recently, an RCT compared the early application of 48 hours of helmet NIV or HFNO in 109 patients with moderate to severe hypoxemia (ratio of partial pressure of arterial oxygen to fraction of inspired oxygen (PaO_2_/FiO_2_) ratio ≤ 200) showed no difference in the number of days free of respiratory support at 28 days (primary outcome) with a significantly lower incidence of intubation and a higher number of invasive mechanical ventilation–free days at 28 days in the helmet NIV group.(8)

The helmet non-invasive ventilation for COVID-19 patients (Helmet-COVID) trial is a concealed, stratified, unblinded multicenter RCT that examines the effect of helmet NIV compared to usual care on 28-day mortality in patients with acute hypoxic respiratory failure due to COVID-19. The full trial protocol has been published previously.(9)

In this manuscript, we describe the statistical analysis plan (SAP) of the helmet COVID trial. This plan complies with the International Conference on Harmonisation of Technical Requirements for Registration of Pharmaceuticals for Human Use, and both the “Statistical principles for clinical trials E9” and “Structure and content of clinical study reports E3”.(10, 11) The report describes the procedures for the primary and secondary analyses. All analyses were prospectively defined as the SAP was finalized during trial implementation. The SAP was written by the Principal Investigator and members of the Steering committee, who will remain blinded to the study results until all patients have been recruited and the database has been locked. Participant recruitment is expected to be complete in the summer of 2021. The final study report will follow the CONSORT (Consolidated Standards of Reporting Trials) 2010 guidelines for reporting RCTs.(12, 13)

## Methods

### Study Design

The Helmet-COVID trial will enroll 320 critically ill patients in Saudi Arabia and Kuwait. The study has been approved by the Institutional Review Boards of the participating sites. The trial is registered in clinicaltrials.gov (NCT04477668). The study is sponsored by King Abdullah International Medical Research Center (Protocol number RC20/306/R), Riyadh, Saudi Arabia. The sponsor has no role in the study design, management, or analysis.

### Study Population

All adult (adult ICU cut-off age) patients admitted to the ICU with suspected or confirmed COVID-19 by reverse transcription–polymerase chain reaction (RT-PCR) will be screened for eligibility. Inclusion criteria include acute hypoxemic respiratory failure (the ratio of arterial oxygen partial pressure to fraction (percent) of inspired oxygen (PaO_2_/FiO_2_ ratio) <200 despite supplemental oxygen with a partial/non-rebreathing mask at a flow rate >10 L/min or above) with intact airway protective gag reflex and able to follow instructions. Exclusion criteria include imminent intubation and requirement for more than one vasopressor. A full list of inclusion and exclusion criteria is described in the published protocol.(9)

A flow diagram will be constructed according to the CONSORT guidelines (Figure 1). We will report the number of patients who were screened, met inclusion or exclusion criteria, were eligible but not enrolled with reasons for non-enrollment. We will report the number of patients who were randomized to each group, received the allocated intervention, and included in the final analysis.

**Figure 1:**
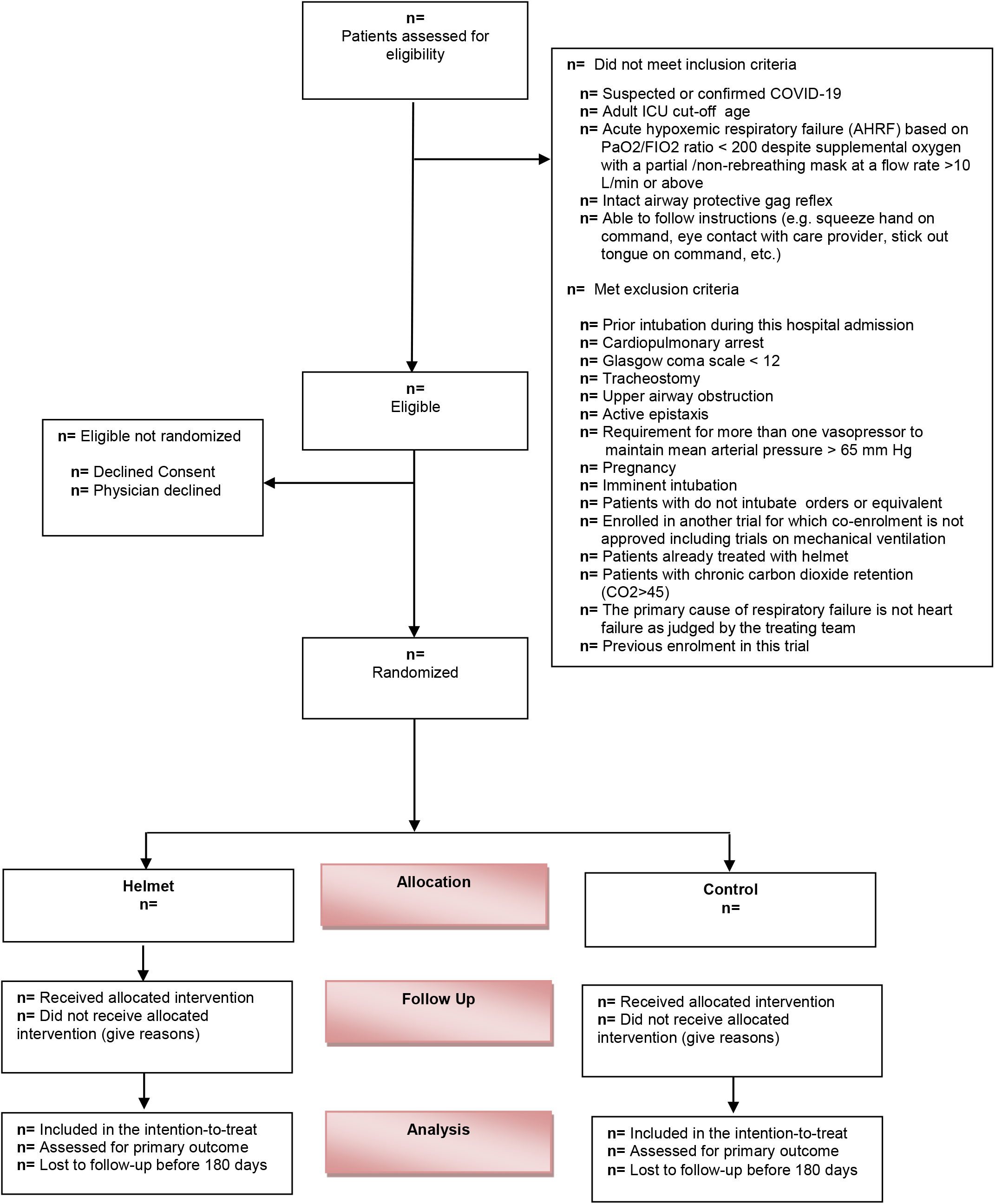
CONSORT Flow Diagram.

*The Intention-to-treat population* consists of all enrolled patients and will be used for the primary analysis. All enrolled patients will be included regardless of whether they receive or do not receive the allocated intervention. All patients enrolled with suspected COVID-19 will remain in the study, even if they tested negative for COVID-19 after enrollment. Post-enrollment exclusion from the intention-to-treat analysis will be restricted to the withdrawal of consent to use trial data by the patient or surrogate decision-maker (SDM) or wrong randomization. However, if the patient or SDM withdraws consent for trial participation but permits collection and use of data, we will include these participants in the planned intention-to-treat analysis. We plan to enroll additional patients to compensate for patients who are excluded post-randomization.

*The per-protocol population* consists of all randomized patients who received the allocated intervention (helmet NIV in the helmet NIV group, and no helmet NIV in the usual care group). Patients will be considered to have received helmet NIV if the device was applied for 1 hour or more.

### Patient and public involvement

No patient involved

## Analysis Plan

### Baseline Characteristics

We will present baseline characteristics in patients randomized to the helmet NIV and usual care of the intention-to-treat cohort (Supplement Table S1). We will report in the two groups patients age, sex, height, weight, body mass index, location before ICU admission (emergency room, hospital ward, other hospitals (ICU or ward), others), Acute Physiology and Chronic Health Evaluation (APACHE) II, sequential organ failure assessment (SOFA) score, comorbidities (any chronic comorbidity, chronic cardiac, pulmonary disease, renal, liver, and neurological diseases, diabetes, any malignancy including leukemia or lymphoma and metastatic solid tumor, AIDS/HIV, rheumatologic diseases, others). Because we are enrolling patients with confirmed or suspected SARS-CoV-2 infection, we will report whether the patient is eventually confirmed to have SARS-CoV-2 infection. We will report physiologic parameters before randomization (partial pressure of oxygen (PaO_2_), fraction of inspired oxygen (FiO_2_), PaO_2_/FiO_2_ ratio, partial pressure of carbon dioxide (PaCO_2_), and pH) and the number of quadrants with infiltrates on the chest radiograph. We will report respiratory support at baseline (high-flow nasal cannula, mask noninvasive ventilation, other oxygen devices). We will document respiratory rate and whether the patient is treated with awake prone positioning. We will document the number of days from onset of symptoms to the emergency room and ICU admission, and the number of days from ICU admission to randomization. We will report organ support, including vasopressor therapy and renal replacement therapy.

### Intervention Data

For each day in the first 96 hours, we will report in each group the details regarding helmet NIV (number of hours used, highest pressure support level, and PEEP). Throughout the first 28 days, we will document the number of days received helmet (>1 hour) and the total hours of helmet NIV. Non-tolerance to helmet NIV is defined as not using helmet NIV for at least 1 hour. We will document the reasons for discontinuation of the helmet NIV (clinical improvement, the patient required intubation, helmet intolerance with use of <1 hour or >1 hour, helmet removal due to change in goals of care, death while on helmet). We will document respiratory support post helmet NIV (mask NIV, high-flow nasal oxygen, other oxygen devices, intubation). Violations to the study protocol will be documented including the use of NIV helmet in the usual group, and lack of attempt to use helmet NIV in the intervention group (Supplement Table S2).

### Co-Interventions

In both groups, we will document the use of other respiratory support modalities during the first 4 days (mask NIV with highest pressure support and PEEP, high-flow nasal cannula with flow rate, other oxygen devices, awake prone position), arterial blood gases, and fluid intake and output (Supplement Table S2).

### Physiologic variables during the intervention

For the 28 days, we will document modalities of respiratory support, the use of sedation while not intubated (dexmedetomidine), renal replacement therapy, and vasopressors/Inotrope therapy. We will document the use of COVID-19 interventions, including corticosteroids, IL-6 receptor antagonists, and antiviral therapy. We will document serial PaO_2_/FiO_2_ ratio, PaCO2, fluid balance, and serial SOFA scores (Supplement Table S2).

### Primary Outcome

The primary outcome is all-cause 28-day mortality. The primary outcome tests the primary hypothesis that helmet NIV reduces 28-day mortality (Supplement Table S3).

### Secondary outcomes

A detailed list of secondary outcomes with definitions has already been published and is outlined in Supplement Table S4. These secondary outcomes can be grouped as follows:

1. Mortality outcomes
  a. ICU mortality
  b. Hospital mortality
  c. 180-day mortality (follow-up study)
2. ICU-free days at day 28
  a. Hospital LOS
  b. Mechanical ventilation free days at day 28
  c. Renal replacement therapy-free days at day 28
  d. Vasopressor-free days at day 28
3. Endotracheal intubation. We will document time to intubation, reasons for intubation as per the treating team (neurologic deterioration that is not attributed to sedation, persistent or worsening respiratory failure of NIV such as oxygen saturation <88%, respiratory rate >36/min, PaO_2_/FiO_2_ ratio <100 or persistent requirement of FiO2 ≥70%, intolerance of mask or helmet NIV, airway bleeding, copious respiratory secretions, respiratory acidosis with pH <7.25, hemodynamic instability, significant radiological worsening). We will document mechanical ventilation parameters in the first 24 hours of intubation (peak airway pressure, plateau pressure, positive end-expiratory pressure (PEEP) (cmH_2_O), FiO_2_ (%), tidal volume, and respiratory rate. We will also document oxygen rescue therapies during invasive mechanical ventilation (neuromuscular blocker infusion, recruitment maneuvers, inhaled nitric oxide, prone positioning, extracorporeal membrane oxygenation (ECMO)). We will report the percentage of patients who underwent tracheostomy.
4. Safety outcomes
  a. Skin injury at the nose, face, neck, and axillae, with the highest stage during the intervention period. We will sue the stages as per the National Pressure Ulcer Advisory Panel(14): stage I: non-blanchable erythema, stage II: partial thickness, stage III: full-thickness skin loss, stage IV: full-thickness tissue loss.
  b. Barotrauma, including pneumothorax, mediastinal air, subcutaneous emphysema.
  c. Cardiovascular events.
  d. Device complications (such as helmet deflation or malfunction).
  e. Serious adverse events (SAEs).

### Statistical analysis

Categorical variables will be reported as numbers and frequencies and will be compared between the study groups using the Chi-square test. Continuous variables will be reported as mean and standard deviation or median and the first and third quartiles (Q1-Q3) and be compared between the study groups using the Student’s t-test or the Wilcoxon-Mann-Whitney test, as judged appropriate by normality testing. For serial measurements, we will test the change over time and the difference between the two study groups over time using a repeated-measures analysis of variance, with no imputation for missing values. For serial measurements, we will use Bonferroni correction to account for multiple comparisons. We will report associations as risk ratios (RR) or hazard ratios (HR) with 95% confidence intervals (CI). Tests will be two-sided and at the 5% significance level. All statistical analyses will be conducted using the SAS software version 9.4 (SAS Institute, Cary, NC, USA). The statistical analysis remains blinded to the research team until completion of primary outcome data on the study population and will be performed by the study biostatistician. A summary of the analysis plan is provided in Table 1.

**Table 1:**
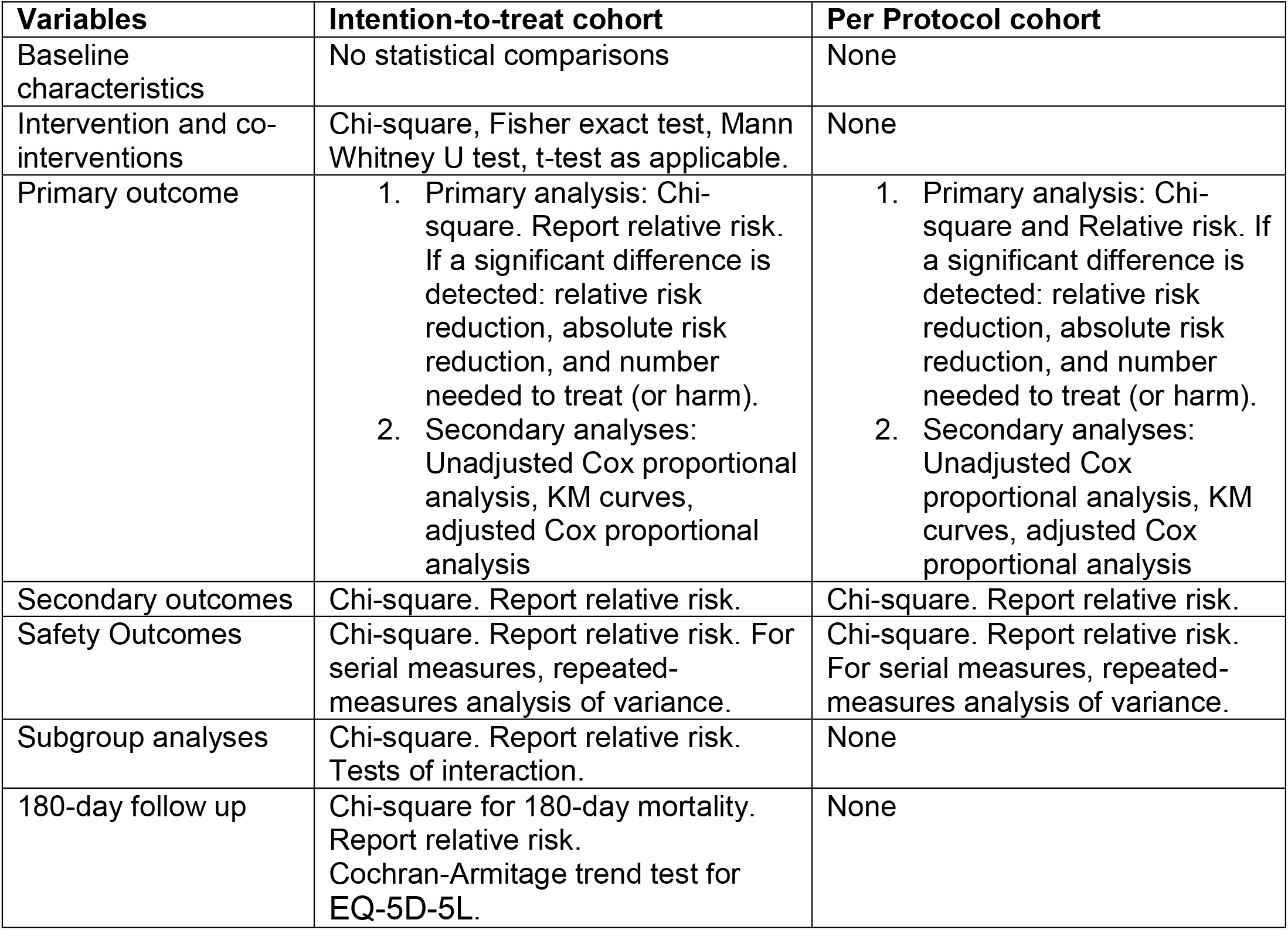
Summary of the analysis plan

| **Variables** | **Intention-to-treat cohort** | **Per Protocol cohort** |
| --- | --- | --- |
| Baseline characteristics | No statistical comparisons | None |
| Intervention and co-interventions | Chi-square, Fisher exact test, Mann Whitney U test, t-test as applicable. | None |
| Primary outcome | 1. Primary analysis: Chi-square. Report relative risk. If a significant difference is detected: relative risk reduction, absolute risk reduction, and number needed to treat (or harm).<br>2. Secondary analyses: Unadjusted Cox proportional analysis, KM curves, adjusted Cox proportional analysis | 1. Primary analysis: Chi-square and Relative risk. If a significant difference is detected: relative risk reduction, absolute risk reduction, and number needed to treat (or harm).<br>2. Secondary analyses: Unadjusted Cox proportional analysis, KM curves, adjusted Cox proportional analysis |
| Secondary outcomes | Chi-square. Report relative risk. | Chi-square. Report relative risk. |
| Safety Outcomes | Chi-square. Report relative risk. For serial measures, repeated-measures analysis of variance. | Chi-square. Report relative risk. For serial measures, repeated-measures analysis of variance. |
| Subgroup analyses | Chi-square. Report relative risk. Tests of interaction. | None |
| 180-day follow up | Chi-square for 180-day mortality. Report relative risk. Cochran-Armitage trend test for EQ-5D-5L. | None |

#### A. Analysis of Primary outcome

The primary outcome will be compared in the intention-to-treat and per-protocol cohorts (effectiveness analysis) using a Chi-square test. Results will be reported as RR with 95% CI. If there is a significant difference, we will report relative risk reduction (RRR), absolute risk reduction (ARR) or increase, and number needed to treat (or harm). The unadjusted cox-proportional hazard model will be used as a secondary analysis tool. Kaplan-Meier curves will be generated for the alternative treatment groups and a log-rank test will be used to compare distributions. Although imbalances in baseline characteristics are unlikely with the large sample size, we will conduct an adjusted logistic regression model to adjust for the following factors (defined a priori): enrolment center and respiratory support at baseline (mask NIV support versus others).

#### B. Analysis of secondary outcomes

Secondary outcomes will be compared in the intention-to-treat cohort only using a Chi-square test. Results will be reported using RR and 95% CI.

#### C. Subgroup analyses

The primary outcome will be compared in the intention-to-treat cohort only, in the following a priori defined subgroups using a Chi-square test.

1. Patients with moderate ARDS (PaO_2_/FIO_2_ ratio 101-200) and patients with severe ARDS (PaO_2_/FIO_2_ ratio <100)
2. Obese patients (body mass index >30 kg/m^2^) and patients with a body mass index of ≤30
3. Patients aged >65 years and ≤65 years
4. APACHE II score higher or lower than the median of enrolled patients
5. Patients who were at the time of enrollment on mask NIV versus others
6. Results will be reported using RR and 95% CI. We will report the results of the test of interactions for these subgroups (Supplement Table S5 online).

#### D. Sensitivity analyses

If patients with suspected COVID-19 who tested negative constituted more than 5% of the study population, we will carry a sensitivity analysis excluding these patients.

#### E. 180-day follow up

In a follow-up report, we will compare 180-day mortality and the EuroQol (EQ-5D-5L)(15) at 6 months scores after randomization between the two study groups.

#### F. Interim analyses

The interim test statistics will be conducted for the primary outcome. We will perform two formal interim analyses when 33% and 67% of the sample size has been reached. The trial may be stopped for safety (p<0.01) or effectiveness (p<0.001) but there will be no plans to terminate the trial for futility. We will account for alpha spending by the O’Brien-Fleming method and the final p-value will be considered at 0.048.(16)

We will also report protocol violations (Supplement Table S6).

## Discussion

Several studies have investigated helmet NIV as a treatment for acute hypoxic respiratory failure.(2-6) A systematic review of RCTs and observational studies found that helmet NIV was associated with lower hospital mortality (odds ratio, 0.43; 95% CI, 0.26-0.69), intubation rate (odds ratio, 0.32; 95% CI, 0.21-0.47), and complications (odds ratio, 0.6; 95% CI, 0.4- 0.92) compared with controls.(17) A meta-analysis of four RCTs (377 patients) showed that helmet NIV significantly increased the PaO_2_/FiO_2_ (+73.4; 95% CI, 43.9-102.9) and decreased the arterial carbon dioxide levels, intubation rate (relative risk, 0.21; 95% CI, 0.11- 0.40) and in-hospital mortality rate (relative risk, 0.22; 95% CI, 0.09-0.50) compared to standard oxygen therapy.(18) A network meta-analysis of 25 studies that included 3804 patients with acute hypoxemic respiratory failure for reasons other than COVID-19 found significantly lower intubation risks with helmet NIV compared with standard oxygen therapy.(7) The advantages of helmet NIV over other oxygen support modalities are thought to be more prominent in patients with COVID-19. This led to the design and conduct of multiple RCTs.(8, 19) Recently, one trial (n=110) showed no difference in the number of days free of respiratory support at 28 days (primary outcome) between helmet NIV and high-flow nasal oxygen.(8)

As the efficacy of helmet NIV to improve outcomes in severe acute hypoxemic respiratory failure due to COVID-19 pneumonia has not been established, the aim of the Helmet COVID trial is to compare the effectiveness of helmet NIV compared to usual care on 28 day - mortality in patients with acute hypoxic respiratory failure from COVID-19. The first patient was enrolled in February 8, 2021. As of May 6, 2021, 108 patients have been enrolled from 4 centers in Saudi Arabia.

## Conclusion

The Helmet-COVID trial evaluates whether helmet NIV improves the outcomes of critically ill patients with acute hypoxemic respiratory failure due to COVID-19. It is expected to provide evidence that will inform practice regarding the use of helmet NIV for respiratory support in these patients and contribute to future clinical practice guidelines.

## Data Availability

Once all planned analyses have been completed and published or presented, data will be shared upon reasonable request from the Chief Investigator.

## List of abbreviations

CI: Confidence Interval
HR: Hazard Ratio
ICU: Intensive Care Unit
IQR: Interquartile Range
LOS: Length of stay
RT-PCR: Reverse Transcription–Polymerase Chain Reaction
RCT: Randomized Controlled Trial
RR: Relative Risk
RRR: Relative Risk Reduction
SAP: Statistical Analysis Plan

## Authors’ contributions

All authors agree to be accountable for all aspects of the work in ensuring that questions related to the accuracy or integrity of any part of the article are appropriately investigated and resolved

## Acknowledgments

The authors would like to thank all the participating patients and their families, as well as the members of the Data Safety & Monitoring Board: Chair: Dr. Nicholas S. Hill (Professor of Medicine, Chief of Pulmonary, Critical Care and Sleep Division, Tufts Medical Center, Boston, Massachusetts, USA), DSM members: Dr. Stefano Nava (Professor of Respiratory Medicine University of Bologna, Chief of the Respiratory and Critical Care Sant’ Orsola Hospital Bologna, Specialist in Respiratory Medicine and Intensive Care Medicine, University of Bologna, Italy), Dr. James Mojica (Vice Chief and Clinical Director of Pulmonary & Critical Care, Director, The Sleep Center at Spaulding, Massachusetts General Hospital, USA) and Dr. Michael Harhay (Assistant Professor of Epidemiology and Medicine-Pulmonary and Critical Care, Department of Biostatistics, Epidemiology and Informatics, University of Pennsylvania USA).

