## Supplementary file for "Statistical Analysis Plan for the Helmet Non-Invasive Ventilation for COVID-19 Patients (Helmet-COVID) Randomized Controlled Trial"

Appendix 1. This appendix was part of the submitted manuscript and has been peer reviewed. It is posted as supplied by the authors

**Table S1: Baseline characteristics – ITT Population.**

| **Characteristic** | **Helmet NIV (N=XXX)** | **Usual Care (N=XXX)** |
| --- | --- | --- |
| Age (Years) - Mean (SD) | xx (xx.x) | xx (xx.x) |
| Male sex - n (%) | xxx/xxx (xx.x) | xxx/xxx (xx.x) |
| Height (cm) - Median (Q1,Q3) | xx (xx.x) | xx (xx.x) |
| Weight (kg) - Median (Q1,Q3) | xx (xx, xx) | xx (xx, xx) |
| BMI (kg/m^2^) - Median (Q1,Q3) | xx (xx, xx) | xx (xx, xx) |
| Location prior to ICU admission - n (%) |  |  |
| Emergency room | xxx (xx.x) | xxx (xx.x) |
| Hospital ward | xxx (xx.x) | xxx (xx.x) |
| Other hospital (ICU or ward) | xxx (xx.x) | xxx (xx.x) |
| Other | xxx (xx.x) | xxx (xx.x) |
| APACHE II- Median (Q1,Q3) | xx (xx, xx) | xx (xx, xx) |
| SOFA score- Median (Q1,Q3) | xx (xx, xx) | xx (xx, xx) |
| Comorbidities - n (%) |  |  |
| Any chronic comorbidity | xxx (xx.x) | xxx (xx.x) |
| Chronic cardiac disease | xxx (xx.x) | xxx (xx.x) |
| Chronic pulmonary disease | xxx (xx.x) | xxx (xx.x) |
| Chronic renal disease | xxx (xx.x) | xxx (xx.x) |
| Mild, moderate or severe liver disease | xxx (xx.x) | xxx (xx.x) |
| Chronic neurological disease, hemiplegia or paraplegia, or dementia | xxx (xx.x) | xxx (xx.x) |
| Diabetes | xxx (xx.x) | xxx (xx.x) |
| Any malignancy including leukemia or lymphoma and metastatic solid tumor | xxx (xx.x) | xxx (xx.x) |
| AIDS/HIV | xxx (xx.x) | xxx (xx.x) |
| Rheumatologic diseases | xxx (xx.x) | xxx (xx.x) |
| Others | xxx (xx.x) | xxx (xx.x) |
| Confirmed SARS-CoV-2 infection* n/N (%) | xxx/xxx (xx.x) | xxx/xxx (xx.x) |
| Physiologic parameters prior to randomization–Median (Q1,Q3) |  |  |
| PaO_2_ (mmHg) | xxx (xx.x) | xxx (xx.x) |
| FiO_2_ | xxx (xx.x) | xxx (xx.x) |
| PaO_2_/FiO_2_ ratio | xxx (xx.x) | xxx (xx.x) |
| PCO_2_ (mmHg) | xxx (xx.x) | xxx (xx.x) |
| HCO_3_ | xxx (xx.x) | xxx (xx.x) |
| pH | xxx (xx.x) | xxx (xx.x) |
| Number of quadrants with infiltrates on chest radiograph | xxx (xx.x) | xxx (xx.x) |
| Respiratory support at baseline- n (%) |  |  |
| High flow nasal cannula | xxx/xxx (xx.x) | xxx/xxx (xx.x) |
| Noninvasive ventilation | xxx/xxx (xx.x) | xxx/xxx (xx.x) |
| Others | xxx/xxx (xx.x) | xxx/xxx (xx.x) |
| Respiratory rate (breaths/minute) –Median (Q1,Q3) | xxx (xx.x) | xxx (xx.x) |
| Awake proning- n (%) | xxx/xxx (xx.x) | xxx/xxx (xx.x) |
| Days from onset of symptoms to the emergency room— Median (Q1, Q3) | xxxx (xx.x) | xxxx (xx.x) |
| Days from onset of symptoms to ICU admission — Median (Q1, Q3) | xxxx (xx.x) | xxxx (xx.x) |
| Number of days from ICU admission to randomization — Median (Q1, Q3) | xx (xx, xx) | xx (xx, xx) |
| Organ support - n (%) |  |  |
| Vasopressors | xxx/xxx (xx.x) | xxx/xxx (xx.x) |
| Renal replacement therapy | xxx/xxx (xx.x) | xxx/xxx (xx.x) |

Denominator of the percentage is the total number of subjects in each group in the ITT population.

For continuous variables, t-test or Mann-Whitney U test will be used to calculate the P value. For categorical variables, chi-square or Fisher’s exact test will be used to calculate the P value unless otherwise noted.

ITT: Intention-to-treat, APACHE: Acute physiology and chronic health evaluation II, Intensive care unit, FiO_2_: denotes the fraction of inspired oxygen, PaO_2_: partial pressure of oxygen in arterial blood, PaCO_2_: partial pressure of carbon dioxide, SOFA: Sequential organ failure assessment.

*Infection is confirmed by a respiratory tract polymerase-chain-reaction test.

**Table S2:** Summary of interventions and co-interventions in the intention-to-treat population.

| **Variable** | **Helmet NIV (N=XXX)** | **Usual Care (N=XXX)** |
| --- | --- | --- |
| Use of helmet |  |  |
| Number of patients receiving helmet during the study period – no. (%) | xxx (xx.x) | xxx (xx.x) |
| Helmet use - Median (Q1, Q3) | xx (xx, xx) | xx (xx, xx) |
| Day 1 number of hours used | xx (xx, xx) | xx (xx, xx) |
| Highest pressure support level | xx (xx, xx) | xx (xx, xx) |
| Highest PEEP | xx (xx, xx) | xx (xx, xx) |
| Day 2 number of hours used |  |  |
| Highest pressure support level | xx (xx, xx) | xx (xx, xx) |
| Highest PEEP | xx (xx, xx) | xx (xx, xx) |
| Day 3 number of hours used |  |  |
| Highest pressure support level | xx (xx, xx) | xx (xx, xx) |
| Highest PEEP | xx (xx, xx) | xx (xx, xx) |
| Day 4 number of hours used |  |  |
| Highest pressure support level | xx (xx, xx) | xx (xx, xx) |
| Highest PEEP | xx (xx, xx) | xx (xx, xx) |
| Number of days received helmet (>1 hour) - Median (Q1, Q3) | xx (xx, xx) | xx (xx, xx) |
| Total hours of helmet | xx (xx, xx) | xx (xx, xx) |
| Reasons for discontinuation of helmet– no. (%) | xxx (xx.x) | xxx (xx.x) |
| Clinical improvement | xxx (xx.x) | xxx (xx.x) |
| Patient required intubation | xxx (xx.x) | xxx (xx.x) |
| Intolerance and helmet use <1 hour |  |  |
| Intolerance and helmet use >1 hour | xxx (xx.x) | xxx (xx.x) |
| Helmet removal due to change in goals of care | xxx (xx.x) | xxx (xx.x) |
| Death while helmet was on | xxx (xx.x) | xxx (xx.x) |
| Respiratory support post helmet – no. (%) |  |  |
| Non helmet noninvasive ventilation | xxx (xx.x) | xxx (xx.x) |
| High flow nasal cannula | xxx (xx.x) | xxx (xx.x) |
| Other oxygen device | xxx (xx.x) | xxx (xx.x) |
| Intubation | xxx (xx.x) | xxx (xx.x) |
| Other respiratory support during the first 4 days |  |  |
| Mask NIV– no. (%) | xxx (xx.x) | xxx (xx.x) |
| Highest pressure support level on day 1 - Median (Q1, Q3) | xx (xx, xx) | xx (xx, xx) |
| Highest PEEP on day 1 - Median (Q1, Q3) | xx (xx, xx) | xx (xx, xx) |
| High flow nasal cannula – no. (%) | xxx (xx.x) | xxx (xx.x) |
| Flow rate on day 1 - Median (Q1, Q3) | xx (xx, xx) | xx (xx, xx) |
| Other oxygen devices | xxx (xx.x) | xxx (xx.x) |
| Awake proning – no. (%) | xxx (xx.x) | xxx (xx.x) |
| Day 1 |  |  |
| FiO2 corresponding to SpO2 | xx (xx, xx) | xx (xx, xx) |
| SpO2 | xx (xx, xx) | xx (xx, xx) |
| S/F ratio | xx (xx, xx) | xx (xx, xx) |
| Fluid intake | xx (xx, xx) | xx (xx, xx) |
| Fluid output | xx (xx, xx) | xx (xx, xx) |
| Day 2 |  |  |
| FiO2 corresponding to SpO2 | xx (xx, xx) | xx (xx, xx) |
| SpO2 | xx (xx, xx) | xx (xx, xx) |
| S/F ratio | xx (xx, xx) | xx (xx, xx) |
| Fluid intake | xx (xx, xx) | xx (xx, xx) |
| Fluid output | xx (xx, xx) | xx (xx, xx) |
| Day 3 |  |  |
| FiO2 corresponding to SpO2 | xx (xx, xx) | xx (xx, xx) |
| SpO2 | xx (xx, xx) | xx (xx, xx) |
| S/F ratio | xx (xx, xx) | xx (xx, xx) |
| Fluid intake | xx (xx, xx) | xx (xx, xx) |
| Fluid output | xx (xx, xx) | xx (xx, xx) |
| Day 4 |  |  |
| FiO2 corresponding to SpO2 | xx (xx, xx) | xx (xx, xx) |
| SpO2 | xx (xx, xx) | xx (xx, xx) |
| S/F ratio | xx (xx, xx) | xx (xx, xx) |
| Fluid intake | xx (xx, xx) | xx (xx, xx) |
| Fluid output | xx (xx, xx) | xx (xx, xx) |
| Co-Interventions – no. (%) |  |  |
| Use of sedation (dexmedetomidine) during NIV support | xxx (xx.x) | xxx (xx.x) |
| Renal replacement therapy | xxx (xx.x) | xxx (xx.x) |
| Vasopressors/Inotropes | xxx (xx.x) | xxx (xx.x) |
| COVID-19 interventions – no. (%) |  |  |
| Hydroxychloroquine | xxx (xx.x) | xxx (xx.x) |
| Chloroquine | xxx (xx.x) | xxx (xx.x) |
| Macrolide | xxx (xx.x) | xxx (xx.x) |
| Lopinavir/Ritonavir | xxx (xx.x) | xxx (xx.x) |
| Favipiravir | xxx (xx.x) | xxx (xx.x) |
| Remdesivir | xxx (xx.x) | xxx (xx.x) |
| Ribavirin | xxx (xx.x) | xxx (xx.x) |
| Intravenous immunoglobulin | xxx (xx.x) | xxx (xx.x) |
| Interferon | xxx (xx.x) | xxx (xx.x) |
| Oseltamivir | xxx (xx.x) | xxx (xx.x) |
| Beta-lactam/Beta-lactamase inhibitor | xxx (xx.x) | xxx (xx.x) |
| Tocilizumab | xxx (xx.x) | xxx (xx.x) |
| Convalescent plasma | xxx (xx.x) | xxx (xx.x) |
| Steroids | xxx (xx.x) | xxx (xx.x) |
| Hydrocortisone | xxx (xx.x) | xxx (xx.x) |
| Dexamethasone | xxx (xx.x) | xxx (xx.x) |
| Prednisolone | xxx (xx.x) | xxx (xx.x) |
| Methyprednisolone | xxx (xx.x) | xxx (xx.x) |
| Other treatments | xxx (xx.x) | xxx (xx.x) |

Calculations are provided for the all patients in each group.

CRRT: continuous renal replacement therapy, FiO_2_: denotes the fraction of inspired oxygen, NIV: non-invasive ventilation, PaO_2_: partial pressure of oxygen in arterial blood, PaCO_2_: partial pressure of carbon dioxide, PEEP: Positive end-expiratory pressure, SOFA: Sequential organ failure assessment.

**Table S3:** Primary Outcome: 28-day mortality

|  | **Intention-to-treat Population** | | | **Per-protocol Population** | | |
| --- | --- | --- | --- | --- | --- | --- |
| **Variable** | **Helmet NIV (N=XXX)** | **Usual Care (N=XXX)** | **P-value** | **Helmet NIV (N=XXX)** | **Usual Care (N=XXX)** | **P-value** |
| 28-day mortality, n (%) | xx/xxx (xx.x) | xx/xxx (xx.x) | x.xxx | xx/xxx (xx.x) | xx/xxx (xx.x) | x.xxx |
| Relative Risk, (95% CI) | xx.x (xx.x, xx.x) | |  | xx.x (xx.x, xx.x) | |  |
| Days to event - Median (Q1,Q3) | xx (xx, xx) | xx (xx, xx) |  | xx (xx, xx) | xx (xx, xx) |  |
| Unadjusted hazard Ratio (95% CI) | x.xx (x.xx, x.xx) | | x.xxx | x.xx (x.xx, x.xx) | | x.xxx |
| Adjusted relative risk, (95% CI) | x.xx (x.xx, x.xx) | | x.xxx | x.xx (x.xx, x.xx) | | x.xxx |

**Table S4:** Secondary and safety outcomes ITT Population

| **Variable** | **Helmet NIV (N=XXX)** | **Usual Care (N=XXX)** | **Relative Risk, (95% CI)** | **P-value** |
| --- | --- | --- | --- | --- |
| 28-day mortality – n (%) | xxxx (xx.x) | xxxx (xx.x) | x.xx (x.xx , x.xx) | x.xxx |
| ICU mortality – n (%) | xxxx (xx.x) | xxxx (xx.x) | x.xx (x.xx , x.xx) | x.xxx |
| Hospital mortality – n (%) | xxxx (xx.x) | xxxx (xx.x) | x.xx (x.xx , x.xx) | x.xxx |
| 180-day mortality – n (%) | xxxx (xx.x) | xxxx (xx.x) | x.xx (x.xx , x.xx) | x.xxx |
| ICU-free days at day 28 | xx (xx, xx) | xx (xx, xx) | x.xx (x.xx , x.xx) | x.xxx |
| Hospital LOS - Median (Q1,Q3) | xx (xx, xx) | xx (xx, xx) | x.xx (x.xx , x.xx) | x.xxx |
| Mechanical Ventilation free days - Median (Q1,Q3) | xx (xx, xx) | xx (xx, xx) | x.xx (x.xx , x.xx) | x.xxx |
| Renal replacement-free days at day 28 | xx (xx, xx) | xx (xx, xx) | x.xx (x.xx , x.xx) | x.xxx |
| Vasopressor-free days at day 28 | xx (xx, xx) | xx (xx, xx) | x.xx (x.xx , x.xx) | x.xxx |
| Endotracheal intubation – n (%) | xxx (xx.x) | xxx (xx.x) | x.xx (x.xx , x.xx) | x.xxx |
| Time to intubation - Median (Q1,Q3) | xx (xx, xx) | xx (xx, xx) | x.xx (x.xx , x.xx) | x.xxx |
| Reasons for intubation – n (%) |  |  |  |  |
| Neurologic deterioration (not attributed to sedation) | xxx (xx.x) | xxx (xx.x) | x.xx (x.xx , x.xx) | x.xxx |
| Persistent or worsening respiratory failure of NIV | xxx (xx.x) | xxx (xx.x) | x.xx (x.xx , x.xx) | x.xxx |
| Oxygen saturation <88% | xxx (xx.x) | xxx (xx.x) | x.xx (x.xx , x.xx) | x.xxx |
| Respiratory rate >36/min | xxx (xx.x) | xxx (xx.x) | x.xx (x.xx , x.xx) | x.xxx |
| P/F ratio <100 | xxx (xx.x) | xxx (xx.x) | x.xx (x.xx , x.xx) | x.xxx |
| Persistent requirement of FiO2 ≥70% | xxx (xx.x) | xxx (xx.x) | x.xx (x.xx , x.xx) | x.xxx |
| Intolerance of face mask or helmet | xxx (xx.x) | xxx (xx.x) | x.xx (x.xx , x.xx) | x.xxx |
| Airway bleeding | xxx (xx.x) | xxx (xx.x) | x.xx (x.xx , x.xx) | x.xxx |
| Copious respiratory secretions | xxx (xx.x) | xxx (xx.x) | x.xx (x.xx , x.xx) | x.xxx |
| Respiratory acidosis with pH <7.25 | xxx (xx.x) | xxx (xx.x) | x.xx (x.xx , x.xx) | x.xxx |
| Hemodynamic instability | xxx (xx.x) | xxx (xx.x) | x.xx (x.xx , x.xx) | x.xxx |
| Significant radiologic worsening | xxx (xx.x) | xxx (xx.x) | x.xx (x.xx , x.xx) | x.xxx |
| Mechanical ventilation parameters in the first 24 hours of intubation |  |  |  |  |
| Ppeak pressure (cmH_2_O) - Median (Q1,Q3) | xx (xx, xx) | xx (xx, xx) |  | x.xxx |
| Pplateau (if done) - Median (Q1,Q3) | xx (xx, xx) | xx (xx, xx) |  | x.xxx |
| PEEP (cmH_2_O) - Median (Q1,Q3) | xx (xx, xx) | xx (xx, xx) |  | x.xxx |
| FiO_2_ (%)- Median (Q1,Q3) | xx (xx, xx) | xx (xx, xx) |  | x.xxx |
| Tidal volume (ml) - Median (Q1,Q3) | xx (xx, xx) | xx (xx, xx) |  | x.xxx |
| Respiratory rate (breaths/min) - Median (Q1,Q3) | xx (xx, xx) | xx (xx, xx) |  | x.xxx |
| Therapies received during invasive mechanical ventilation – n (%) |  |  |  |  |
| Neuromuscular blocker infusion | xxx (xx.x) | xxx (xx.x) | x.xx (x.xx , x.xx) | x.xxx |
| Recruitment maneuvers | xxx (xx.x) | xxx (xx.x) | x.xx (x.xx , x.xx) | x.xxx |
| Inhaled Nitric oxide | xxx (xx.x) | xxx (xx.x) | x.xx (x.xx , x.xx) | x.xxx |
| Prone positioning | xxx (xx.x) | xxx (xx.x) | x.xx (x.xx , x.xx) | x.xxx |
| ECMO | xxx (xx.x) | xxx (xx.x) | x.xx (x.xx , x.xx) | x.xxx |
| Tracheostomy – n (%) | xxx (xx.x) | xxx (xx.x) | x.xx (x.xx , x.xx) | x.xxx |
| Safety outcomes |  |  |  |  |
| Skin ulceration at nose, face, neck and axillae (highest stage during intervention period) |  |  |  |  |
| Stage I: Non-blanchable erythema | xxx (xx.x) | xxx (xx.x) | x.xx (x.xx , x.xx) | x.xxx |
| Stage II: Partial thickness | xxx (xx.x) | xxx (xx.x) | x.xx (x.xx , x.xx) | x.xxx |
| Stage III: Full thickness skin loss | xxx (xx.x) | xxx (xx.x) | x.xx (x.xx , x.xx) | x.xxx |
| Stage IV: Full thickness tissue loss | xxx (xx.x) | xxx (xx.x) | x.xx (x.xx , x.xx) | x.xxx |
| Barotrauma | xxx (xx.x) | xxx (xx.x) | x.xx (x.xx , x.xx) | x.xxx |
| Pneumothorax | xxx (xx.x) | xxx (xx.x) | x.xx (x.xx , x.xx) | x.xxx |
| Mediastinal air | xxx (xx.x) | xxx (xx.x) | x.xx (x.xx , x.xx) | x.xxx |
| Subcutaenous emphysema | xxx (xx.x) | xxx (xx.x) | x.xx (x.xx , x.xx) | x.xxx |
| Cardiovascular events | xxx (xx.x) | xxx (xx.x) | x.xx (x.xx , x.xx) | x.xxx |
| Device complication (helmet deflation) | xxx (xx.x) | xxx (xx.x) | x.xx (x.xx , x.xx) | x.xxx |
| Serious adverse events (SAEs) | xxx (xx.x) | xxx (xx.x) | x.xx (x.xx , x.xx) | x.xxx |
| SAE 1 | xxx (xx.x) | xxx (xx.x) | x.xx (x.xx , x.xx) | x.xxx |
| SAE 2 | xxx (xx.x) | xxx (xx.x) | x.xx (x.xx , x.xx) | x.xxx |
| SAE 3 | xxx (xx.x) | xxx (xx.x) | x.xx (x.xx , x.xx) | x.xxx |

Denominator of the percentage is the total number of subjects in each group in the ITT and PP population.

Mechanical Ventilation free days Vasopressor free days and ICU free days are calculated based on 28-d observation

LOS: Length of Stay, ECMO: Extracorporeal membrane oxygenation; ICU: Intensive care unit

For continuous variables, the Mann-Whitney U test was used to calculate p value

For categorical variables, the Fishers exact test was used to calculate p value

**Table S5:** Subgroup analyses

|  | **28-day mortality** | | | | |
| --- | --- | --- | --- | --- | --- |
|  | **Helmet NIV (N=XXX)** | **Usual Care (N=XXX)** | **RR (95% CI)** | **P-value** | **P-value for interaction** |
| Moderate ARDS | xxx/xxx (xx.x) | xxx/xxx (xx.x) | x.xx (x.xx, x.xx) | x.xxx | x.xxx |
| Severe ARDS | xxx/xxx (xx.x) | xxx/xxx (xx.x) | x.xx (x.xx, x.xx) | x.xxx |  |
| BMI >30 | xxx/xxx (xx.x) | xxx/xxx (xx.x) | x.xx (x.xx, x.xx) | x.xxx | x.xxx |
| BMI ≤30 | xxx/xxx (xx.x) | xxx/xxx (xx.x) | x.xx (x.xx, x.xx) | x.xxx |  |
| Age >65 | xxx/xxx (xx.x) | xxx/xxx (xx.x) | x.xx (x.xx, x.xx) | x.xxx | x.xxx |
| Age ≤65 | xxx/xxx (xx.x) | xxx/xxx (xx.x) | x.xx (x.xx, x.xx) | x.xxx |  |
| APACHE II >Median | xxx/xxx (xx.x) | xxx/xxx (xx.x) | x.xx (x.xx, x.xx) | x.xxx | x.xxx |
| APACHE II ≤Median | xxx/xxx (xx.x) | xxx/xxx (xx.x) | x.xx (x.xx, x.xx) | x.xxx |  |

ARDS: Acute respiratory distress syndrome, BMI: Body mass index, APACHE: Acute physiology and chronic health evaluation II

**Table S6:** Summary of Protocol Violations

| **Characteristics** | **Helmet NIV (N=XXX)** | **Usual Care (N=XXX)** |
| --- | --- | --- |
| Protocol violations | xx (xx.x) | xx (xx.x) |
| xxxxxxx | xx (xx.x) | xx (xx.x) |
| xxxxxxx | xx (xx.x) | xx (xx.x) |
| Reasons for protocol violations |  |  |
| xxxxxxx | xx (xx.x) | xx (xx.x) |
| xxxxxxx | xx (xx.x) | xx (xx.x) |
| Any consequences |  |  |
| Yes | xx (xx.x) | xx (xx.x) |
| No | xx (xx.x) | xx (xx.x) |
| Denominator of the percentage is the total number of patients in the treatment group. | | |

**Table S7:** EQ-5D-5L Frequencies and proportions at baseline and day 180 post randomization

|  | **Baseline** | | | **180-day** | | |
| --- | --- | --- | --- | --- | --- | --- |
| Variable | **Helmet NIV (N=XXX)** | **Usual Care (N=XXX)** | **P value** | **Helmet NIV (N=XXX)** | **Usual Care (N=XXX)** | **P value** |
| Mobility |  |  |  |  |  |  |
| I have no problems with walking around |  |  |  |  |  |  |
| I have slight problems with walking around |  |  |  |  |  |  |
| I have moderate problems with walking around |  |  |  |  |  |  |
| I have severe problems with walking around |  |  |  |  |  |  |
| I am unable to walk around |  |  |  |  |  |  |
| Self-care |  |  |  |  |  |  |
| I have no problems with washing or dressing myself |  |  |  |  |  |  |
| I have slight problems with washing or dressing myself |  |  |  |  |  |  |
| I have moderate problems with washing or dressing myself |  |  |  |  |  |  |
| I have severe problems with washing or dressing myself |  |  |  |  |  |  |
| I am unable to wash or dress myself |  |  |  |  |  |  |
| Usual Activities |  |  |  |  |  |  |
| I have no problems doing my usual activities |  |  |  |  |  |  |
| I have slight problems doing my usual activities |  |  |  |  |  |  |
| I have moderate problems doing my usual activities |  |  |  |  |  |  |
| I have severe problems doing my usual activities |  |  |  |  |  |  |
| I am unable to do my usual activities |  |  |  |  |  |  |
| Pain Discomfort |  |  |  |  |  |  |
| I have no pain or discomfort |  |  |  |  |  |  |
| I have slight pain or discomfort |  |  |  |  |  |  |
| I have moderate pain or discomfort |  |  |  |  |  |  |
| I have severe pain or discomfort |  |  |  |  |  |  |
| I have extreme pain or discomfort |  |  |  |  |  |  |
| Anxiety Depression |  |  |  |  |  |  |
| I am not anxious or depressed |  |  |  |  |  |  |
| I am slightly anxious or depressed |  |  |  |  |  |  |
| I am moderately anxious or depressed |  |  |  |  |  |  |
| I am severely anxious or depressed |  |  |  |  |  |  |
| I am extremely anxious or depressed |  |  |  |  |  |  |

Cochran-Armitage trend test is used to calculate the p value

Figures

**Figure 1:** Kaplan Meier curve for mortality

**Figure 2:** Kaplan Meier curve for time to intubation

**Figure 3:** Serial SOFA, Serial P/F ratios, Serial PaCO_2_

**Figure 4:** Visual Analog Scale – Dyspnea, Visual Analog Scale – Device discomfort

**Figure 5:** EQ-5D-5L at baseline and at Day 180 (follow-up study)
